# Women’s views on accepting COVID-19 vaccination during and after pregnancy, and for their babies: A multi-methods study in the UK

**DOI:** 10.1101/2021.04.30.21256240

**Authors:** Helen Skirrow, Sara Barnett, Sadie Bell, Lucia Riaposova, Sandra Mounier-Jack, Beate Kampmann, Beth Holder

**Author notes:** Corresponding author: Dr. Beth Holder, Imperial College London, Institute of Reproductive and Developmental Biology, Hammersmith Campus, London W12 0HS. These authors contributed equally (Helen Skirrow, Sara Barnett).

## Abstract

**Background:** COVID-19 vaccines are the cornerstone of the pandemic response and now advised for pregnant women in the United Kingdom(UK) however COVID-19 vaccine acceptance among pregnant women is unknown.

**Methods:** An online survey and semi-structured interviews were used to investigate pregnant women’s views on COVID-19 vaccine acceptability for themselves when pregnant, not pregnant and for their babies. 1,181 women, aged over 16 years, who had been pregnant since 23rd March 2020, were surveyed between 3^rd^ August–11^th^ October 2020. Ten women were interviewed.

**Results:** The majority of women surveyed (81.2%) reported that they would ‘definitely’ or were ‘leaning towards’ accepting a COVID-19 vaccine when not pregnant. COVID-19 vaccine acceptance was significantly lower during pregnancy (62.1%, p<0.005) and for their babies (69.9%, p<0.005). Ethnic minority women were twice as likely to reject a COVID-19 vaccine for themselves when not pregnant, pregnant and for their babies compared to women from White ethnic groups (p<0.005). Women from lower-income households, aged under 25-years, and from some geographic regions were more likely to reject a COVID-19 vaccine when not pregnant, pregnant and for their babies. Multivariate analysis revealed that income and ethnicity were the main drivers of the observed age and regional differences. Women unvaccinated against pertussis in pregnancy were over four times more likely to reject COVID-19 vaccines when not pregnant, pregnant and for their babies. Thematic analysis of the survey freetext responses and interviews found safety concerns about COVID-19 vaccines were common though wider mistrust in vaccines was also expressed. Trust in vaccines and the health system were also reasons women gave for accepting COVID-19 vaccines.

**Conclusion:** Safety information on COVID-19 vaccines must be clearly communicated to pregnant women to provide reassurance and facilitate informed pregnancy vaccine decisions. Targeted interventions to promote COVID-19 vaccine uptake among ethnic minority and lower-income women may be needed.

## Background

On the 16^th^ April 2021, the United Kingdom’s (UK) Joint Committee on Vaccination and Immunisation (JCVI) announced that pregnant women should be offered the COVID-19 vaccine *‘at the same time as the rest of the population, based on their age and clinical risk group’* (1). Given this latest guidance change, understanding pregnant women’s’ perspectives on the acceptability of being vaccinated against COVID-19 is vital. We present here the first multi-methods study exploring UK women’s views on the acceptability of COVID-19 vaccination in pregnancy, as well as their views on vaccination for their babies, and for themselves when not pregnant.

At the start of the SARS-CoV-2 pandemic in 2020, there was a lack of evidence on the risk of COVID-19 disease in pregnant women (2). It is now known that while pregnant women do not appear to be at greater risk of contracting SARS-CoV-2 there is a small risk of severe illness with COVID-19 disease, particularly in the last trimester of pregnancy (i.e. from 28 to 40 weeks of pregnancy) (2-4). Since the beginning of the pandemic in the United Kingdom (UK), as a precaution pregnant women have been classed as ‘vulnerable’ to COVID-19 and advised to carefully adhere to social distancing guidance by the National Health Service (NHS), particularly in the third trimester of pregnancy (5, 6).

Despite calls by experts (7) pregnant women were not included in the initial COVID-19 vaccine trials, though COVID-19 vaccine trials involving pregnant women have now started (8). In the UK, the general COVID-19 vaccination programme began in December 2020 with prioritisation of those at greater risk of hospitalisation or being severely ill with COVID-19 and those caring for vulnerable individuals, such as health and social care workers (9). Initial guidance from the UK’s Joint Committee on Vaccination and Immunisation (JCVI), was that pregnant women should not be offered COVID-19 vaccination due to a lack of data on the safety of COVID-19 vaccines during pregnancy (9). In contrast, in the United States of America (USA) the Centers for Disease Control and Prevention (CDC) advised that pregnant women could be offered COVID-19 vaccination with information available to enable pregnant women to make informed decisions (10). The UK guidance was changed on December 30^th^ 2020 (11), with pregnant women at greater risk of contracting COVID-19 (e.g. frontline healthcare workers) or at greater risk of severe disease due to other risk factors being able to be vaccinated following a discussion with a healthcare professional (11). Given the availability of a larger databases on vaccine safety following the introduction of the vaccines, primarily from the USA (12), this recommendation was recently further amended to include all pregnant women in line with the rest of the population, with mRNA vaccines identified as the preferred product to be offered (1).

Ethnic minorities are at higher risk of dying from COVID-19 (13), and in the UK pregnant women from Black or other ethnic minority groups are overrepresented among women admitted to hospital with COVID-19 infection during pregnancy (4). Recent work by Bell et al found that parents from ethnic minority backgrounds other than White in the UK are less likely to accept a COVID-19 vaccine for their children (14). This finding is consistent with other reports that individuals from ethnic minorities are less likely to accept COVID-19 vaccination for themselves (15, 16). For example, initial analysis of the UK’s COVID-19 vaccine program for adults aged over 80 years found lower uptake among Black Afro-Caribbean ethnicities (17). We have previously shown that acceptance of pertussis and influenza vaccines in pregnancy is also lower in this group (18, 19).

Parental decisions about childhood vaccinations have also been shown to begin in pregnancy (20), so it is therefore useful to assess pregnant women’s perspectives on COVID-19 vaccines for both themselves and their children. Understanding women’s views and acceptability of COVID-19 vaccination is also important to address circulating misinformation about COVID-19 vaccines impacting on fertility (21) given that exposure to misinformation about COVID-19 vaccines can influence vaccine intentions (22).

We conducted a multi-methods study to investigate the views of pregnant women in the UK on the likely uptake of a future COVID-19 vaccine for themselves and their children. At the time of the survey no COVID-19 vaccines had been licensed for use but there was a recognition that COVID-19 vaccination could be made routinely available to pregnant women, children, and women of childbearing age in the future.

## Methods

A multi-methods approach was taken – using quantitative and qualitative components – with the aim of gaining insight into the acceptability of future COVID-19 vaccines for pregnant women and their children at a time when no COVID-19 vaccines had yet been licensed for use. The data presented here is part of a larger survey aimed at investigating the impact of the COVID-19 pandemic on access, awareness, and acceptance of routine maternal vaccines. The study comprised of a questionnaire survey and semi-structured interviews, to both quantify different views on accepting COVID-19 vaccines and then to also explore the reasons for these views in more depth.

### Survey Recruitment

Eligible participants were required to have been pregnant at some point between the start of the UK 2020 lockdown (from 23rd March 2020) and the time of survey completion, to be resident in the UK, and to be aged 16 years or over. The survey was live from 3^rd^ August – 11^th^ October 2020. The online survey was prefaced by an information page explaining the study, and how the data was to be used. Participants were informed that by taking part in the survey, they agreed for their responses to be used for research purposes. Participants were required to confirm (by tick-box) at the start of the survey that they met the eligibility criteria and that they consented to participate in the survey.

The survey was advertised and promoted using Facebook with a Facebook landing page and paid advertising using Facebook’s ad manager which cross posts to Instagram. The three adverts had a combined reach of 46,146, 1,573 post engagements and 1,394 link clicks. Related organisations on Facebook were also contacted individually by study researchers, including pregnancy yoga and birth preparation classes, breastfeeding support groups and toddler groups. The survey was shared and distributed via the research team’s personal twitter accounts including linking to other researchers and organisations with maternal and vaccine uptake interests. Finally, the survey was also promoted via some Maternity Voices Partnerships (23) who were e-mailed and invited to share the survey, and via a post on the website Mumsnet.

### Survey design

The survey was designed with input from midwives, pregnancy vaccine researchers, paediatricians and public health professionals and was based on previous research surveys on pregnancy vaccination (18) and other surveys that had been used to assess COVID-19 vaccine views during the pandemic (14). Here we present one aspect of the survey, specifically regarding the acceptability of a ‘future’ COVID-19 vaccination. The COVID-19 vaccine section asked respondents: ‘*Please select how much you agree or disagree with the following statements about a future vaccine to protect against COVID-19: i) If a vaccine against coronavirus (COVID-19) becomes available, I would get vaccinated* ***whilst pregnant***, *ii) If a vaccine against coronavirus (COVID-19) becomes available, I would get vaccinated* ***whilst not pregnant*** *and iii) If a vaccine against coronavirus (COVID-19) becomes available, I would vaccinate* ***my baby***.*’* Responses were scored on a Likert scale: *‘Yes definitely’, ‘Unsure but leaning towards yes’, ‘Unsure but leaning towards no’, ‘No, definitely not’*. This question was followed by a free-text box titled: *‘Feel free to add any additional comments here’*.

This anonymous survey gathered optional demographic data including ethnicity, age, number of children, country of residence, region of residence in England, ethnicity, parity, income, pregnancy status, gestation at survey completion for those who were pregnant and date of delivery for those who had already had their babies. At the end of the online survey, participants were invited to take part in a follow-up interview by leaving their contact details; they were informed that by leaving their details, their responses would no longer be anonymous.

### Survey analysis

Responses were scored on a Likert scale coded as follows: 1) *‘Yes definitely’, 2) ‘Unsure but leaning towards yes’, 3) ‘Unsure but leaning towards no’, 4) ‘No, definitely not’*. Acceptability of a COVID-19 vaccine for women when pregnant, when not pregnant and for their child was compared in all survey respondents by Pearson’s chi-square followed by analysis of each cell’s contribution to the chi squared statistic. Ordered logistic regressions of the Likert responses were conducted to determine the demographic factors associated with maternal acceptability of the COVID-19 vaccine for themselves whilst pregnant and not pregnant, and for their child. Ethnicity, age, country of residence, region of residence (for residents in England) and household income, were analysed separately and then in multivariate models to determine predictors of acceptability across the UK and within England. Odds ratios over 1 represent an increased likelihood of women’s responses moving from ‘yes, definitely’ towards ‘definitely not, and odds ratios of less than 1 represent a movement in the opposite direction. Some groups were combined as follows. 1) The <20y group was combined with the next age bracket to form < 25y age bracket. The ten income groups were combined pair-wise to form five groups. To enable multiparametric analysis, ethnicity was dichotomised into ‘White’ (i.e., White British, White Irish and White Other participants) and ‘Ethnic minorities’ (i.e., Black, Asian, Chinese, Mixed ethnicities or Other ethnicity). A p value of less than 0.05 was considered statistically significant. Sankey diagrams created using SankeyMATIC.

### Semi-structured interviews

Participants who had left their contact details at the end of the online survey were contacted by SBa. Participants were purposively selected to prioritise respondents who; 1) were from ethnic minority backgrounds, due to lower representation among survey respondents; 2) were pregnant at the time of survey completion, due to their proximity to their pregnancy experience compared to those that had already had their babies at the time of survey completion; 3) had not completed the open text survey responses. Informed consent was obtained by telephone or e-mail, depending on participant preference. The participant information sheet and consent form were provided by e-mail (see supplementary material A). Interviews lasted approximately 30 minutes and were conducted over the telephone and/or using Microsoft Teams, and were recorded with permission of the participant. Interviews were conducted by SBa and HS using a topic guide. The topic guide was developed based on the questionnaire (see supplementary material B). The data presented here pertains to their responses regarding views on accepting the COVID-19 vaccine when pregnant, not pregnant and for their children. The interviews took place between 7th-16th December 2020.

### Qualitative analysis

Free-text responses following the survey questions on COVID-19 vaccine acceptance were analysed thematically by SBa using the stages outlined by Braun and Clarke: data familiarisation, coding and theme identification and refinement (24). To enhance the rigour of the analysis, coding approaches and subsequent theme generation and refinement was discussed between HS, SBa, SBe and BH. Interviews were transcribed verbatim and analysed thematically following a similar approach as the free-text survey responses, initially by SBa and then with agreement by SBa, SBe HS and BH.

### Ethical approval

This study was approved by Imperial College Research Ethics Committee (ICREC) (Ref: 20IC6188).

## Results

### Demographics of survey respondents

There were 1,526 respondents to the survey in total. 120 responses were excluded because they were test responses or incomplete responses, leaving 1,406 responses. Of these, there were 1,181 respondents to the questions regarding acceptability of COVID-19 vaccination. Demographic details, for those respondents that chose to provide these, are summarised in Figure 1. The most common age group was 30-34 years (n=461), and the majority of respondents were White British (n=1092). Most women worked full-time (n=739), and median household annual income was £45,999-54,999. The majority of respondents were from England (n=1083) though a higher proportion of respondents were from London and the South-West regions of England (see figure 1F). 68% of women were pregnant at time of survey completion, whilst the remaining 32% had given birth. Those who were pregnant ranged from between 5 to 41 weeks gestation, with 34 weeks being the most frequent gestation (n=50 - see figure 1H). Most women responding either had no other children (n=450) or one child (n=430).

**Figure 1:**
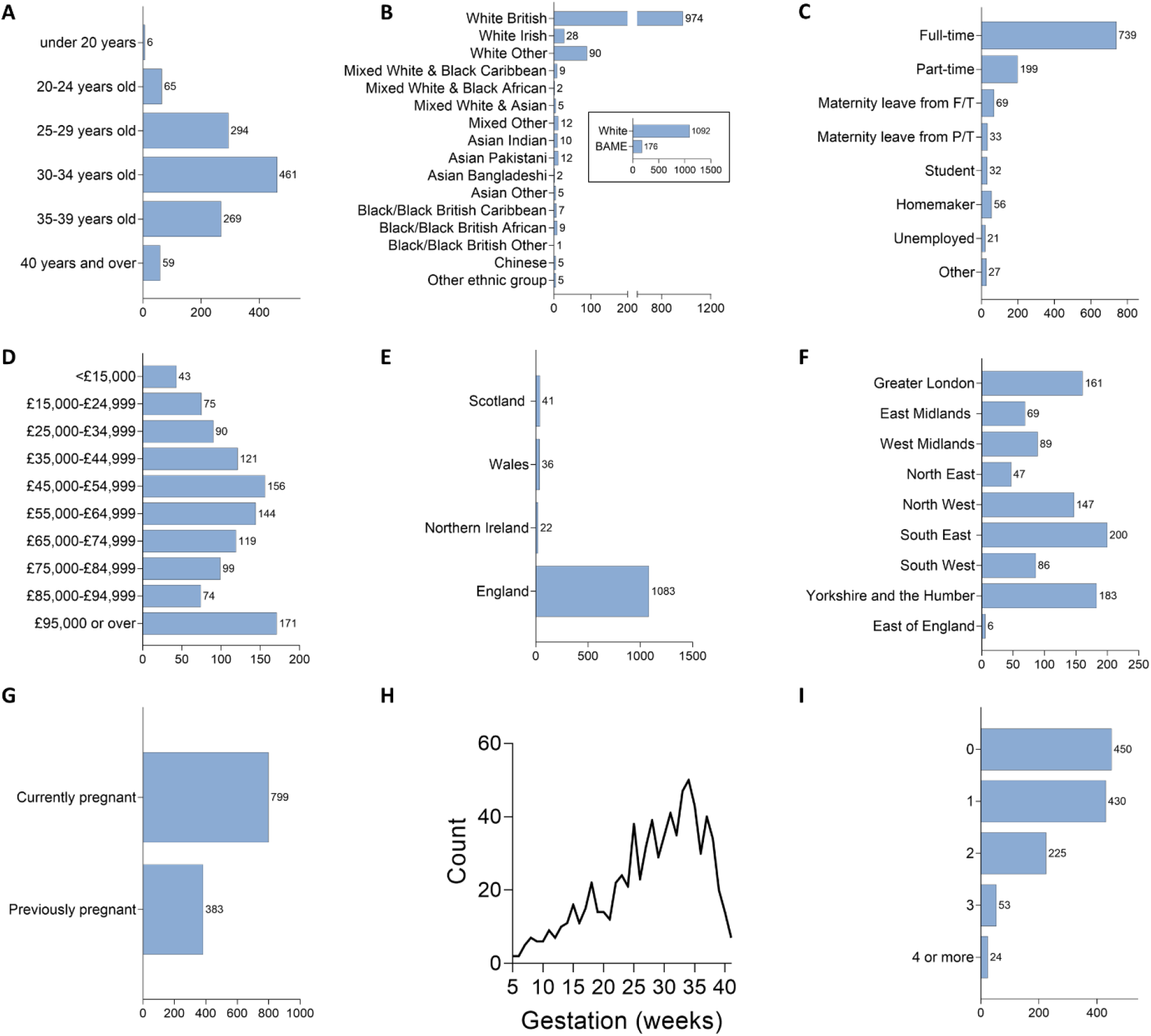
Demographics of Survey Respondents. Self-reported demographics of the survey respondents at the time of survey completion. These questions were optional. A) Age (years); B) Ethnicity; C) Employment status; D) Annual household income (£); E) Country of residence; F) Region of residence for English residents (n=988); G) Pregnancy status; H) Gestational age of pregnancy (weeks) for those currently pregnant; I) Number of children.

### Acceptability of a future COVID-19 vaccination

The majority (81.2%) of survey respondents reported that they would ‘definitely’ accept (55.1%), or were ‘leaning towards’ (26.1%) accepting a future COVID-19 vaccine for themselves (Figure 2A). Most (62.1%) respondents also reported that they would definitely accept, or were leaning towards accepting a future COVID-19 vaccine for themselves *whilst pregnant*. Acceptance of vaccination during pregnancy was significantly lower than acceptance outside of pregnancy (p<0.005; Figure 2A). Analysis of the contribution to this chi square result showed that this was mainly driven by a difference in selection of ‘no, definitely not’, with 2.3 times more respondents expressing this perspective regarding vaccination in pregnancy (17.8%), compared to 7.6% regarding vaccination whilst not pregnant. The second driver of this difference was selection of the ‘yes, definitely’ response, with 55.1% of women expressing this perspective regarding COVID-19 vaccination for themselves, which dropped to 21.2% regarding vaccination during pregnancy.

**Figure 2:**
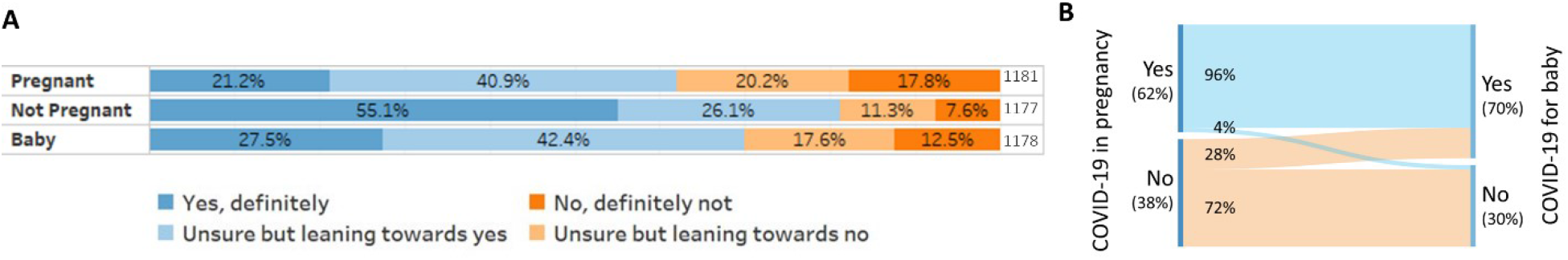
Women’s acceptability of a future COVID-19 vaccine during pregnancy, after pregnancy and for their baby. Survey respondents were asked how much they agreed or disagreed about a future vaccine to protect against COVID-19 for delivery whilst they were Pregnant, whilst Not Pregnant or for their Baby. Responses were scored on a Likert scale (see key). A) All responses to the survey question (n=1177-1181); B) Sankey plot of all respondents showing linkage between their acceptance of a future COVID-19 vaccine during pregnancy and their acceptance of the same vaccine for their babies (n=178).

Most (69.9%) respondents reported that they would definitely accept (27.5%), or were leaning towards (42.4%) accepting a future COVID-19 vaccine for their babies (Figure 2A). This was significantly lower than their acceptability of COVID-19 vaccination for themselves (p<0.005; Figure 2A). Again, this difference was primarily driven by a difference in the selection of ‘no, definitely not’, with 12.5% of women selecting this option for their baby, followed by selection of ‘yes, definitely’, with only 27.5% of women selecting this option for their baby, compared to 55.1% selecting it for themselves. Almost all (96%) of women who reported they were likely to accept COVID-19 vaccination for themselves during pregnancy, also reported they were likely to accept the vaccine for their babies, compared to only 28% of those who reported they were likely to reject the vaccine for themselves (Figure 2B).

### Acceptability of future COVID-19 based on pregnancy status at time of survey completion

Survey respondents who were pregnant when they completed the survey were more likely to reject the idea of receiving a COVID-19 vaccine during pregnancy compared to those who were no longer pregnant when they completed the survey (p<0.005; Figure 3A). 68.1% of those who were no longer pregnant said they would definitely or were leaning towards accepting a future vaccine during pregnancy, compared to 59.2% of those who were still pregnant. There was no difference in acceptability of a vaccine to be delivered outside of pregnancy. Respondents who had already had their baby were also more likely to accept the idea of receiving a COVID-19 vaccine for their babies, compared to those who were still pregnant (p<0.005, Figure 3A).

**Figure 3:**
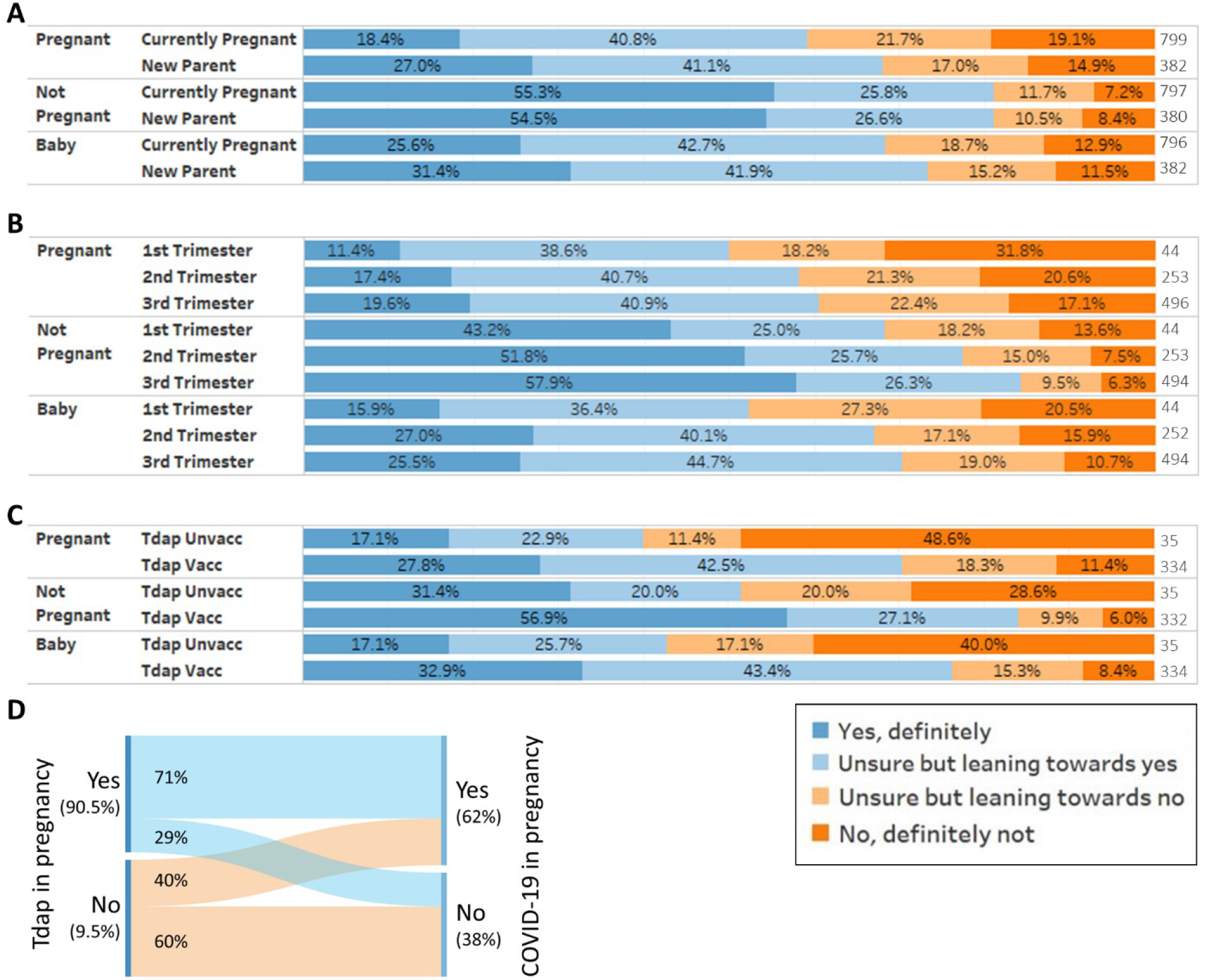
Women’s acceptability of a future COVID-19 vaccine during pregnancy, after pregnancy and for their baby, by pregnancy status. Survey respondents were asked how much they agreed or disagreed about a future vaccine to protect against COVID-19 for delivery whilst they were Pregnant, whilst Not Pregnant or for their Baby. Responses were scored on a Likert scale (see key). A) COVID-19 vaccine acceptance split by whether women were pregnant at the time of the survey (n=796-799), or were new mothers (n=382); B) COVID-19 vaccine acceptance split by gestation at time of survey completion; C) COVID-19 vaccine acceptance in new mothers (n=382) split by whether they had been vaccinated against pertussis (Tdap vacc) or not (Tdap unvacc) during their last pregnancy; D) Sankey plot of new mothers (n=382) showing linkage between Tdap vaccination status in their last pregnancy and their acceptance of a future COVID-19 vaccine in pregnancy.

Of women who were pregnant at time of survey completion, those in the third trimester of pregnancy were more likely to accept the idea of a future COVID-19 vaccine for their baby compared to respondents in the first trimester (p=0.025; Figure 3B). They also had a non-significant higher likelihood of accepting a future COVID-19 vaccine during pregnancy (p=0.057).

### Acceptability of future COVID-19 vaccine based on pertussis vaccine status during pregnancy

Of women who had delivered their babies, those who had not been vaccinated against pertussis in pregnancy were around four times (p value <0.0005) more likely to reject the COVID-19 vaccine during pregnancy, outside of pregnancy and for their baby (Figure 3C). Only 40% of unvaccinated women reported they were likely to accept a future COVID-19 vaccine, compared to 71% of pertussis-vaccinated women (Figure 3D).

### Demographic factors associated with future COVID-19 vaccine acceptability

#### Ethnicity

Compared to women of white ethnicities, women from Black and Mixed Black ethnicity groups (Black-British African, Black-British Caribbean, Black-other, Mixed White-Black Caribbean, and Mixed White-Black African) were more likely to reject a future COVID-19 vaccine when pregnant, not pregnant or for their baby (p<0.005, Figure 4A, Supplementary Table 1). 16.2% respondents of all white ethnicities reported that they would definitely not accept a future COVID-19 vaccine compared to 46.4% of Black and Black Mixed ethnicity respondents (Figure 4A). Similarly, 21.8% of white respondents answered that they would definitely accept a future COVID-19 vaccine compared to 3.6% of Black and Mixed Black ethnicity respondents (Figure 4A). The effect of ethnicity remained significant in multivariate analysis (Table 1).

**Figure 4:**
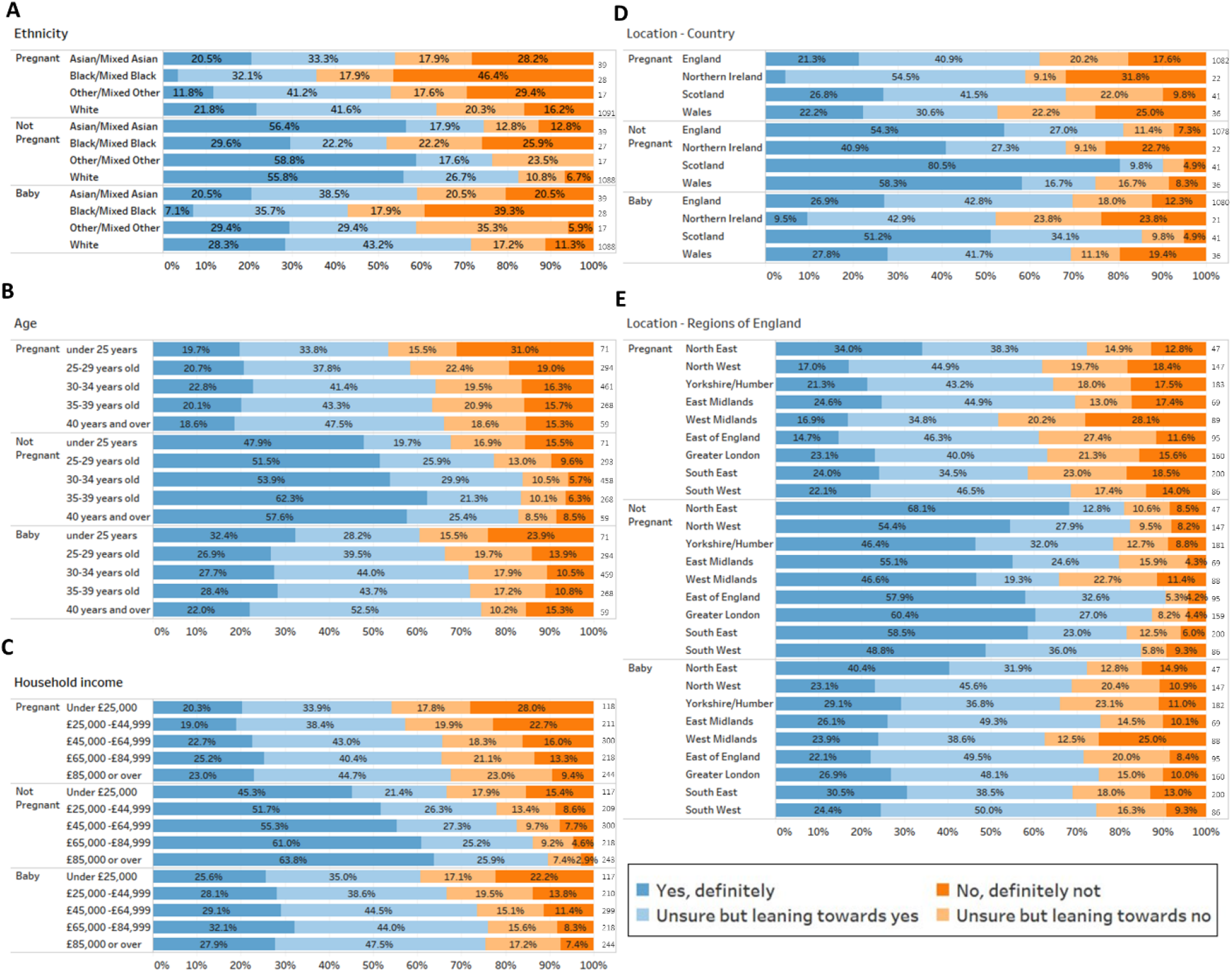
Women’s acceptability of a future COVID-19 vaccine during pregnancy, after pregnancy and for their baby split by respondent demographics. Survey respondents were asked how much they agreed or disagreed about a future vaccine to protect against COVID-19 for delivery whilst they were Pregnant, whilst Not Pregnant or for their Baby. Responses were scored on a Likert scale (see key). Responses are separated by self-reported demographics. These questions were optional, so the number of responses vary. The number in each group is shown to the right of each bar. A) COVID-19 vaccine acceptance split by ethnicity. B) COVID-19 vaccine acceptance split by age; C) COVID-19 vaccine acceptance split by household income; D) COVID-19 vaccine acceptance split by country of residence; E) COVID-19 vaccine acceptance split by region of residence for English residents.

**Table 1:** Multivariate analysis of predictors of COVID-19 vaccine acceptance.

| Variable | Pregnant |  |  | Non-pregnant |  |  | Baby |  |  |
| --- | --- | --- | --- | --- | --- | --- | --- | --- | --- |
|  | OR | 95% CI | P value | OR | 95% CI | P value | OR | 95% CI | P value |
| **Ethnicity** |  |  |  |  |  |  |  |  |  |
| White# | - | - | - | - | - | - | - | - | - |
| Minority ethnicity | 2.20 | 1.45, 3.33 | **<0.005** | 1.86 | 1.21, 2.87 | **0.005** | 2.17 | 1.43, 3.28 | **<0.005** |
| **Age** |  |  |  |  |  |  |  |  |  |
| Under 25y | 1.13 | 0.69, 1.88 | 0.614 | 1.10 | 0.66, 1.84 | 0.706 | 0.89 | 0.53, 1.49 | 0.668 |
| 25 – 29y | 1.11 | 0.84, 1.45 | 0.473 | 1.07 | 0.80, 1.43 | 0.636 | 1.05 | 0.80, 1.39 | 0.712 |
| 30 – 34 y # | - | - | - | - | - | - | - | - | - |
| 35 – 39 y | 1.03 | 0.78, 1.36 | 0.845 | 0.75 | 0.55, 1.03 | 0.075 | 0.92 | 0.69, 1.22 | 0.573 |
| Over 39y | 0.91 | 0.56, 1.49 | 0.713 | 0.82 | 0.48, 1.42 | 0.484 | 0.99 | 0.60, 1.64 | 0.976 |
| **Country** |  |  |  |  |  |  |  |  |  |
| England# | - | - | - | - | - | - | - | - | - |
| Scotland | 0.75 | 0.42, 1.33 | 0.321 | 0.31 | 0.14, 0.68 | **0.004** | 0.37 | 0.20, 0.68 | **0.001** |
| Wales | 1.25 | 0.66, 2.36 | 0.500 | 0.87 | 0.44, 1.74 | 0.436 | 1.02 | 0.53, 1.94 | 0.957 |
| Northern Ireland | 1.73 | 0.80, 3.74 | 0.163 | 2.02 | 0.89, 4.58 | 0.890 | 2.22 | 1.01, 4.85 | **0.046** |
| **Income** |  |  |  |  |  |  |  |  |  |
| Under £24,999 | 1.98 | 1.36, 2.89 | **<0.005** | 2.59 | 1.73, 3.86 | **<0.005** | 2.18 | 1.50, 3.18 | **<0.005** |
| £25,000-£44,999 | 1.53 | 1.08, 2.18 | **0.017** | 1.66 | 1.14, 2.43 | **0.009** | 1.28 | 0.90, 1.82 | 0.174 |
| £45,000-£64,999 | 1.15 | 0.84, 1.58 | 0.384 | 1.42 | 1.00, 2.01 | **0.048** | 1.09 | 0.79, 1.50 | 0.598 |
| £65,000-£84,999 | 1.06 | 0.76, 1.48 | 0.736 | 1.14 | 0.79, 1.66 | 0.484 | 0.94 | 0.67, 1.31 | 0.701 |
| Over £85,000# | - | - | - | - | - | - | - | - | - |

**OR:** ordinal odds ratio. An OR above 1 indicates a higher likelihood of women giving responses moving from 'definitely yes' towards 'definitely no' on the Likert scale. **95% CI:** 95% confidence interval. **#** indicates the comparator group in the analysis. **Ethnicity Groups: White:** White-British, White-Irish, White-Other; **Minority ethnicity:** Black-British African, Black-British Caribbean and Black-Other, Asian Indian, Asian-Pakistani, Asian-Bangladeshi, Asian-Other and Chinese, Mixed White-Black Caribbean, Mixed White-Black African, Mixed White-Asian, Mixed White/Other and Other ethnicity.

#### Age

In univariate analysis, respondents aged under 25 years were more likely to reject a future COVID-19 vaccine whether pregnant or not pregnant, compared to the middle age bracket of 30-34 (p<0.05 –Figure 4B and supplementary Table 1) but were equally accepting of a future COVID-19 vaccine for their baby (p>0.05). In the multivariate analysis, which also took into account ethnicity, location and income, the relationship between younger age and being more likely to reject a future COVID-19 vaccine was no longer significant (Table 1).

#### Income

Compared to the highest income bracket, women with lower annual household incomes were more likely to reject a COVID-19 vaccine for themselves and for their babies (Figure 3C and Supplementary Table 1). Women with household incomes less than £25,000 were 3 times more likely to say definitely no to a COVID-19 vaccine in pregnancy (28%) compared to the highest income bracket (>£80,000; 9.4%). In multivariate analysis, which took into account ethnicity, location and age, lower income remained significantly associated with reduced acceptance (Table 1).

#### Geographical Location – Country of the United Kingdom

Respondents from Scotland were more likely to accept a future COVID-19 vaccine for themselves whilst not pregnant and for their baby compared to respondents from England (p<0.05; Figure 4D, Supplemental Table 1). Almost twice the number of residents in Scotland responded ‘definitely yes’ (51.2%) to a vaccine for their baby compared to English residents (26.9%). These differences remained once ethnicity, income and age were taken into account (Table 1). Similarly, respondents from Northern Ireland were more likely to reject a future COVID-19 vaccine for their babies compared to respondents from England (p=0.032; Figure 4D) which also remained significant in multivariate analysis (see below).

#### Geographical Location – Region of England

In univariate analysis, women located in the West Midlands were more likely to reject a future COVID-19 vaccine for themselves when pregnant (p=0.027), not pregnant (p=0.002) and for their babies (p=0.042; Figure 4E) compared to respondents from Greater London. Respondents from Yorkshire were also more likely to reject a future COVID-19 vaccine when not pregnant (p=0.006; Figure 4E). These regional differences were lost once ethnicity, income and age were taken into account (Table 1). Step-wise inclusion of variables indicated that the lower acceptance in the West Midlands was accounted for by income rather than ethnicity (data not shown).

### Multivariate analysis of demographic factors associated with COVID-19 vaccine acceptability

We performed two multivariate logistic regressions-one for all residents of the UK (Table 1) and one for residents of England (Supplementary Table 2), taking into account ethnicity, age, household income and location. Across the UK, predictors of lower acceptance of a future COVID-19 vaccination when pregnant were being from an ethnic minority (p<0.005), and having a household income lower than £45,000 (p=<0.005 for <£25,000; p=0.017 for £25,000-44,999). For acceptance of a future COVID-19 vaccine when not pregnant, being from an ethnic minority (p=0.005), and having a household income below £65,000 (p=<0.0005 for <£25,000; p=0.009 £25,000-44,999; p=0.048 for £45,000-64,999) were also associated with lower acceptance. In addition, being a resident of Scotland was independently associated with higher acceptance (p=0.004). Predictors of lower acceptance of a future COVID-19 vaccine for respondents’ babies were being from an ethnic minority (p=<0.005) and having a household income below £25,000 (p=<0.005). Being a resident of Scotland was also associated with higher acceptance of a future COVID-19 vaccine for respondents’ babies (p=0.001) while respondents from Northern Ireland were more likely to reject a future COVID-19 vaccine for their baby (p=0.046).

Within England, following inclusion of other variables, the association between women living in the West Midlands being more likely to reject a COVID-19 vaccine became non-significant (p=0.058). Once ethnicity, location and income were taken into account, age was no longer a predictor of vaccine acceptance for any scenario nor living in any region of England. See table 1 for breakdown of results and supplementary table 2.

### Qualitative results

Of the 1,181 respondents to the COVID-19 vaccine acceptance question in the survey, 19.7% (n=233) of respondents left a response in the subsequent free-text box. The number of responses to each theme by survey respondent is shown in Table 2 including how often themes were mentioned by the women surveyed. Semi-structured interviews took place with 10 women who were from 6 different regions of England, the most common area being London where 4 of the women lived. The women interviewed were aged between 25-40 years and further demographics are displayed in Table 3.

**Table 2:**
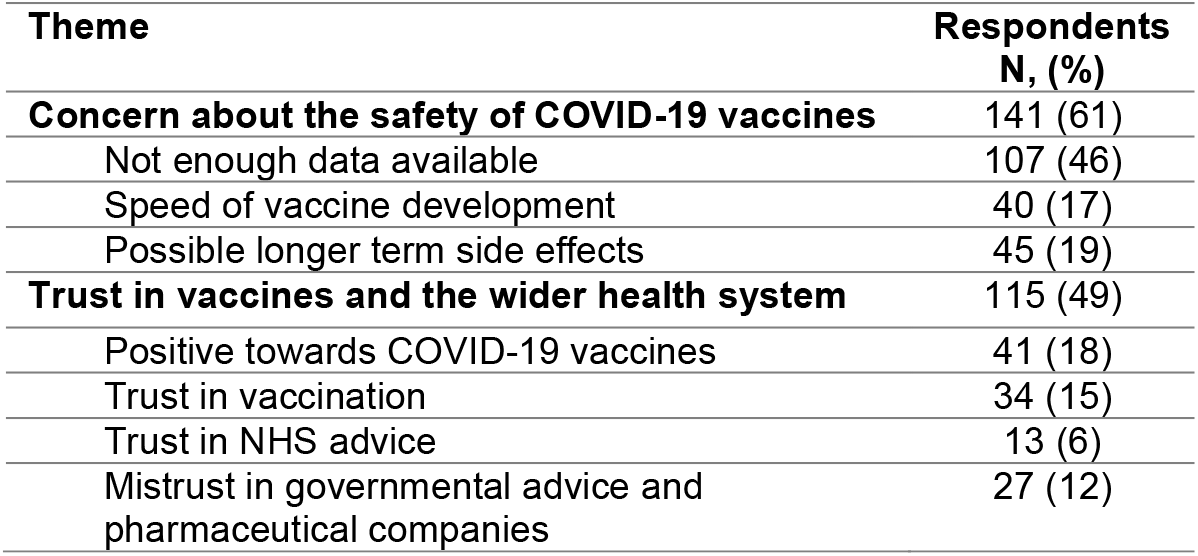
Themes identified from free-text survey and interview responses.

**Table 3:** Interviewee characteristics.

| No | Ethnicity | Parity | Tdap | Would accept COVID-19 vaccine- |  |  |
| --- | --- | --- | --- | --- | --- | --- |
|  |  |  |  | -when pregnant | -when not pregnant | -for baby |
| 1 | Chinese | 3 | Yes | No | No | No |
| 2 | White Asian | 1 | Yes | Yes | Yes | Yes |
| 3 | British Pakistani | 1 | No | No | Yes | No |
| 4 | British Arabic | 1 | Yes | No | Yes | No |
| 5 | Black-African | 1 | Yes | No | No | No |
| 6 | White British | 2 | Yes | No | Yes | No |
| 7 | White British | 4 | Yes | Yes | Yes | Yes |
| 8 | White British | 2 | Yes | No | Yes | No |
| 9 | White British | 2 | Yes | Yes | Yes | Yes |
| 10 | White British | 1 | Yes | No | Yes | No |

Thematic analysis of the freetext survey responses and interviews found that safety concerns (61% of women surveyed who left freetext answers) were the most common reason women gave for influencing whether they would accept a COVID-19 vaccine during pregnancy. Safety concerns were linked to concerns about lack of safety data (46% of women surveyed who left freetext answers) and worries around the speed of vaccine development (17% of women surveyed who left freetext answers). Women also commonly (49% of women surveyed who left freetext answers) mentioned trust or mistrust in wider vaccination and the health system as reasons for declining or accepting future COVID-19 vaccines. Thematic analysis of both the survey responses and interviews are outlined below.

#### • Concern about the safety of COVID-19 vaccines

In free-text responses and the interviews, many women expressed safety concerns, particularly related to feeling that there was ‘***not enough data available on the COVID-19 vaccines’*** for either them or their babies to be vaccinated – a theme also linked to the speed of development of the COVID-19 vaccine. Women expressed that there was *“Not enough evidence to say it would work, no evidence to say it doesn’t cause side effects*.*”(survey participant # S1212)* or that they would like *“to read more evidence/studies and science on vaccinations against Covid before considering my baby having it or myself whilst pregnant*.*” (survey participant # S1275)*. Women particularly wanted data on the safety of the vaccine on foetuses and whilst breastfeeding: *“I’d want reassurance that there was absolute confidence that there were no harmful long-term effects on my baby or no chance of it causing any harm in utero’ (survey participant #s1228) and “I would also want more information on how it would affect my baby if I was breastfeeding, and I received it*.*” (survey participant #s1340)*. One interviewee described that *“once it’s been declared safe for me to have it, I would go ahead and get it done”*(Interviewee 09).

Another common safety concern expressed by women was the **‘*speed of vaccine development’*** of COVID-19 vaccines and therefore the ***‘newness’*** of any future COVID-19 vaccines. For example, one respondent reported: *“I would be very cautious over a new vaccine that has been created in such short space of time when usually they take years*.*” (survey participant # S746)*. Whilst another respondent stated: *“No one in their right mind would accept a vaccine that’s been rushed’ (survey participant # S1497)*. There was a feeling that with time after a COVID-19 vaccine had been widely used that this would increase acceptability amongst women: *“My worry is that it hasn’t had long enough to understand any longer-term effects of the vaccine yet*.*” (survey participant # S1426)*. Another respondent said: *“Vaccines take years to perfect, there is no way I am vaccinating either my baby or myself with a vaccine that has only just been found!” (survey participant #s1236)*. One interviewee said: *“Because it’s really new, there’s not a lot of, um, like there’s not a lot of research into it yet. I know they’ve just taken it out and it’s like a short period of time otherwise vaccines take a good few years before they come out. So, I just think the timeframe and things*.*”* (Interviewee 03).

Many comments reflected worries about the ‘***possible long-term side effects of a Covid - 19 vaccines***’ not having been studied: *“No long-term side effects or risks are known about any vaccine for coronavirus therefore I wouldn’t be comfortable vaccinating myself or my children*.” *(survey participant # S1499)*. This theme was commonly also related to wanting more evidence on safety such as for example: ‘*I have to see what might be the long-term effects because now we don’t know about the long term effects about the vaccine. Maybe it’s now it’s effective, right now, but no one knows what can happen with the people that get the vaccine now*.*”* (Interviewee 05). The concerns about the side effects were also related to the speed of development: *“I feel vaccines might be rushed without thorough long-term side-effect testing. So would want to wait for evidence they are 100% safe*.” (*survey participant # S1425)*.

#### • Trust in vaccines and the wider health system

Women who expressed views that were ‘***positive responses towards COVID-19 vaccines’*** often mentioned this as part of a wider trust and confidence in vaccines and the health system: *“I strongly believe in vaccination. I trust the UK’s method in developing a COVID vaccine and it’s safety*.*” (survey participant # S1457)* and *“I would trust my GP and midwives if they recommended it*.*” (survey participant # S1247)*.

Respondents therefore reported being confident and having ‘***trust in vaccination’*** in general and therefore being willing to accept the COVID-19 vaccine if it was recommended or deemed safe: *“if told safe by researchers/GP, would get vaccinated in pregnancy for Covid 19 definitely*.*”(survey participant # S1317)*.

***‘Trust in NHS advice’*** was also expressed as a reason for accepting a future COVID-19 vaccine *“I also understand that vaccinations for pregnant women and young babies would not be offered on the NHS if they weren’t safe. So, if they were being offered on the NHS then yes, I would have them*.*” (survey participant # S1529)*

Concerns about the speed of the development of the vaccine in the context of the global pandemic also related to ‘***mistrust in government’*** regarding the handling of the COVID-19 pandemic and also ‘**mistrust in wider pharmaceutical industry’**. For example: ***“****I feel it’s so early in understanding the virus to be vaccinated, all of the science has been confusing and changes far too often and doesn’t go far enough to answer why BAME communities are most at risk. I’m not comfortable with the advice being given and the government’s ability to be truthful so I will not be getting a vaccination and I would not want to put my baby at risk either” (S1199 Q55)*

Respondents also expressed mistrust about a COVID-19 vaccine for example: *“I would not trust a vaccine against Covid so I would never get it or my baby. I feel it would be very dangerous to our health*.” *(survey participant # S1270)*. These sentiments were often expressed again as mistrust in vaccine manufacturing in general for example: *“I feel very sceptical and paranoid about whether I was to get vaccinated if a COVID vaccine was available. This is due to scientists disagree with each other and the contradictions presented by vaccine developers” (survey participant # S1368)*.

#### • Themes only identified in interviews

Some additional themes not identified in the free-text responses emerged from the interviews. Interviewees commonly acknowledging ‘**that children are not at increased risk from coronavirus’** which links to the lower likelihood of acceptance of infant vaccination seen in our quantitative analysis (Figure 2A). For example: *“children are less, so are less likely to have corona than maybe grown-ups, so I don’t think I will get my baby the vaccine*.*”* (Interviewee 05). Interviewees also **acknowledged that the COVID-19 vaccine offered a route to resuming normal life**: *“I feel, you know, they’ve passed it, it’s gone through vigorous testing. Yes, it may have been quite quick, but it needed to be quick obviously. I think the more people that are vaccinated, the quicker we may get back to some form of normality, it may not go back to what it was before, but I definitely think it’s a good thing*”(Interviewee 06).

## Discussion

Our survey of 1,181 UK residents found that the majority of women would accept or were leaning towards accepting a COVID-19 vaccine. Acceptability was highest for themselves to be vaccinated when not pregnant, with over 8 in 10 of women answering they would likely accept the COVID-19 vaccine. A significantly lower proportion of 6 in 10 women would accept or were leaning towards accepting a COVID-19 vaccine when pregnant, and fewer women answered that they would definitely accept vaccination when pregnant. However, less than 2 in 10 women said they would definitely not accept a COVID-19 vaccination in pregnancy. The majority of respondents would either accept or were leaning towards accepting a COVID-19 vaccine for their baby, although this was also significantly lower than vaccine acceptance for themselves.

Women belonging to an ethnic minority group were twice as likely to reject a COVID-19 vaccine for themselves when pregnant, not pregnant and for their babies compared to women from White ethnicity groups. Women with lower annual household income were also more likely to reject the COVID-19 under all circumstances. Respondents living in Scotland were more accepting of COVID-19 vaccination, independent of income and ethnicity, with 5 of 10 respondents from Scotland saying they would definitely accept vaccination for their baby compared to only 1 in 10 respondents from Northern Ireland. Women who had not been vaccinated against pertussis in pregnancy were four times more likely to also reject the COVID-19 vaccine independent of pregnancy status or for their baby.

Qualitative analysis of both the survey responses and the semi-structured interviews found that women expressed concerns about the COVID-19 vaccine development as they perceived it as having been rushed and they wanted more information on safety and side effects. It would be of great interest to repeat this survey now that there is more data on these aspects following the publication of data from large COVID-19 vaccine trials in adults, though vaccine trial data on pregnant women and children is still not available. Thematic analysis identified that respondents who trusted vaccination in general expressed confidence in accepting COVID-19 vaccines, and if they were recommended by the NHS for pregnant women. This supports our finding that pertussis vaccination uptake in pregnancy predicted women’s acceptance of future COVID-19 vaccination. Some respondents expressed feelings of mistrust in COVID-19 vaccines, wider vaccination programmes and health system advice, which are areas that should be prioritised for public health messages.

A study by Skjefte et al surveyed COVID-19 vaccine acceptance among 17,871 pregnant women and mothers of children aged under 18 years from sixteen countries, including 2,702 respondents from the United Kingdom (25). They report lower COVID-19 vaccine acceptance among pregnant women from all countries surveyed (52%) compared to our findings though similar COVID-19 vaccine intentions for their children (69.2%,(25)) to our findings. COVID-19 vaccine acceptance among pregnant women therefore varies by country (25) which is also supported by another multi-country survey assessing pregnant women’s attitudes towards COVID-19 vaccines (26). Ceulemans et al, though having fewer UK respondents than our study also found vaccine acceptance was lower among pregnant women compared to non-pregnant women (26). Skjetfte et al also found that confidence in overall vaccination programmes and trust in public health systems were predictors of COVID-19 vaccine acceptance in pregnancy (25), including among UK respondents, which is consistent with our qualitative findings. Another recently published survey of 300 pregnant women in Turkey also found a lower acceptance of COVID-19 vaccine in pregnancy (37%) than our findings (27). Therefore, vaccine programmes must be tailored to reflect individual countries specific contexts and levels of vaccine acceptance.

Our findings are also consistent with other surveys assessing COVID-19 vaccine acceptability outside of pregnancy, which also found that ethnic minorities are less likely to accept vaccination in the UK (14, 16). For example, we recently reported that parents of Black, Asian, Chinese, Mixed or Other ethnicities were almost 3 times more likely to reject a COVID-19 vaccine for themselves and their children compared to White ethnicity parents in England (14). Though the majority of the UK public are vaccine confident (28), COVID-19 vaccine confidence among ethnic minority communities has been a concern (29, 30). Initial analysis of COVID-19 vaccine uptake among over 80 year old’s in the UK in January 2021 Black-Afro Caribbean ethnicity individuals were less likely to have received their COVID-19 vaccine (17). This variation in uptake is a particular worry if it was to correlate to COVID-19 vaccine uptake among pregnant women given the higher COVID-19 pregnancy admissions experienced among women from ethnic minorities in the UK (4).

Routine vaccination in pregnancy is now established worldwide with pregnant women in the United Kingdom offered vaccination against pertussis and seasonal influenza and there are more vaccines on the horizon (31). Uptake of existing maternal vaccines can be affected by both access and vaccine confidence (32) and in the UK women living in poorer areas or belonging to a minority ethnicity group are less likely to be vaccinated in pregnancy (19, 33, 34). This is mirrored in uptake of childhood vaccines in the UK, with children also living in poorer areas or belonging to a minority ethnicity group being less likely to receive their routine vaccinations (33, 35, 36). Our findings suggest that women from lower income households and belonging to ethnic minority groups are less likely to accept COVID-19 vaccines when pregnant, not pregnant and for their baby and therefore reflect existing maternal vaccine uptake inequalities. This is important to address given the recent change to guidance which now recommends that COVID-19 vaccines can be offered to pregnant women in the UK (1). Our findings suggest that regional variation within England, or uptake in lower age groups is driven by lower income and ethnicity. Lower income can intersect with lower education levels (29) and vaccine hesitancy towards COVID-19 vaccines has been found to correlate with lower education levels (37). Therefore tailored, accessible information to overcome hesitancy among some population groups is needed (29). Our survey had lower numbers in some regions, and a larger study may be able to detect additional regional differences to those reported here to further understand variation in uptake now the COVID-19 vaccination programme is established.

Our findings add weight to the calls to involve pregnant women in vaccine trials earlier, in order for vaccine safety data to be available so that pregnant women to make informed vaccine decisions (7, 38, 39). Free-text responses and interview responses both showed that the main concern about COVID-19 vaccines was around safety. There was significant lower acceptance of vaccination in pregnancy, suggesting that women are understandably more cautious about receiving a new vaccine whilst pregnant and they want safety information and data that is directly related to pregnancy. Thus, the earlier availability of vaccine safety data relating to pregnancy, and its communication, is vital. Since our survey and interviews were carried out there has been misinformation around COVID-19 vaccines and women of child-bearing age – including inaccurate rumours around the COVID-19 vaccine impacting on fertility meaning accurate safety communication to all women is needed (21, 40, 41).

Our findings also support previous research relating pregnancy vaccine attitudes to parental vaccine decisions for children (20). We found that women not vaccinated against pertussis in pregnancy were four times more likely to say they would not accept a future COVID-19 vaccine when pregnant, not pregnant and for their baby. Thus, improving vaccine information delivery in pregnancy may also improve subsequent childhood vaccine acceptance and indeed wider COVID-19 vaccine acceptance. Our qualitative findings found that wider mistrust in vaccines and the health system was associated with COVID-19 vaccine concerns. Promoting vaccination in pregnancy (32) potentially therefore offers an opportunity to promote vaccination along the life course.

### Strengths and Limitations

The main strength of this study was the use of multiple methods – the qualitative analysis of the survey and interviews enabled factors behind the quantitative findings to be explored in more detail. The response rate with over 1000 responses from women who had been pregnant during the first peak of the COVID-19 pandemic in the UK was excellent with nearly 1 in 5 respondents leaving freetext responses to the COVID-19 vaccine acceptability questions.

The survey included respondents from across the United Kingdom, which enabled us to identify interesting differences in Scotland and Northern Ireland compared to England, however the majority of respondents were from England. Regionally, London and the South-East were overrepresented, however the survey respondents included women with a range of ages and income levels and at different pregnancy gestations. Importantly, the survey captured women with a range of vaccine attitudes and women who had both been vaccinated with pertussis vaccine in pregnancy and those who had not. Although we were able to detect a significantly lower uptake in some ethnic minority groups (Black ethnicities including Black-British African, Black-British Caribbean, Black-other and Black Mixed ethnicities) we were underpowered to detect differences in other ethnic groups. Thus to explore the role of ethnicity in multivariate analyses we had to combine ethnic groups and ethnicity was dichotomised into White ethnicities and all other ethnic minorities.

The strength of this survey was that it took place in the middle of the pandemic, at a time of great uncertainty and upheaval for both patients and the healthcare system. However, this is also the main limitation of the survey, as when the survey was live between August and October 2020 no COVID-19 vaccine had yet been licensed. The survey may have found different results if it had taken place once the COVID-19 vaccines had been approved in the United Kingdom as studies have found that vaccine attitudes towards the COVID-19 vaccines have changed over time (42). However, despite COVID-19 vaccines now having been approved for pregnant women there remains uncertainty around COVID-19 vaccination in pregnancy due to emerging safety data and changing governmental advice (1, 11). Our findings therefore remain timely and relevant.

### Conclusion

Our findings support the need for clear accurate communication to reassure pregnant women about COVID-19 vaccine safety particularly given that COVID-19 vaccines have now been recommended for pregnant women in the UK. Repeating the survey following the recent guidance change for vaccination in pregnancy may be beneficial to understand current concerns of pregnant women about the COVID-19 vaccines. Monitoring COVID-19 vaccine uptake among different income and ethnicity groups is needed to ensure existing inequalities in vaccine uptake in general and among pregnant women are not exacerbated. Targeted interventions to maximize COVID-19 vaccine uptake among pregnant women from ethnic minority communities and those living in deprived areas should be explored.

## Data Availability

All of the reported data is contained within the manuscript.

## Acknowledgements

We would like to thank all survey and interview participants for their time to provide their views and opinions. Thank you to the Bay Wide, Royal Surrey, Sandwell and West Birmingham, Leicester City, Bradford, Bolton, Central Cheshire, Chester, Bath and North-East Somerset, Swindon and Wiltshire, Royal Free and Kingston Maternity Voices Partnerships who agreed to share the survey via their networks.

Mr Toby Clements and Dr Thomas Rice helped with creation and dissemination of the online survey.

## References

1. JCVI. Joint Committee on Vaccination and Immunisation. JCVI issues new advice on COVID-19 vaccination for pregnant women 16 Apr 2021 [Available from: https://www.gov.uk/government/news/jcvi-issues-new-advice-on-covid-19-vaccination-for-pregnant-women.

2. Mullins E, Evans D, Viner R, O’Brien P, Morris E. Coronavirus in pregnancy and delivery: rapid review. Ultrasound in Obstetrics & Gynecology. 2020;55(5):586–92.

3. Khalil A, Kalafat E, Benlioglu C, O’Brien P, Morris E, Draycott T, et al. SARS-CoV-2 infection in pregnancy: A systematic review and meta-analysis of clinical features and pregnancy outcomes. EClinicalMedicine. 2020;25:100446.

4. Knight M, Bunch K, Vousden N, Morris E, Simpson N, Gale C, et al. Characteristics and outcomes of pregnant women admitted to hospital with confirmed SARS-CoV-2 infection in UK: national population based cohort study. BMJ. 2020;369:m2107.

5. NHS. Pregnancy and coronavirus 2021 [Available from: https://www.nhs.uk/conditions/coronavirus-covid-19/people-at-higher-risk/pregnancy-and-coronavirus/.

6. RCOG. Coronavirus (COVID-19) Infection in Pregnancy 2021 [Available from: https://www.rcog.org.uk/globalassets/documents/guidelines/2021-02-19-coronavirus-covid-19-infection-in-pregnancy-v13.pdf.

7. Heath PT, Le Doare K, Khalil A. Inclusion of pregnant women in COVID-19 vaccine development. The Lancet Infectious Diseases. 2020;20(9):1007–8.

8. Pfizer. NEWS/ Pfizer and BioNTech Commence Global Clinical Trial to Evaluate COVID-19 Vaccine in Pregnant Women 2021 [Available from: https://www.pfizer.com/news/press-release/press-release-detail/pfizer-and-biontech-commence-global-clinical-trial-evaluate.

9. JCVI. Joint Committee on Vaccination and Immunisation: advice on priority groups for COVID-19 vaccination. 2 December 2020 [Available from: https://assets.publishing.service.gov.uk/government/uploads/system/uploads/attachment_data/file/948353/Priority_groups_for_coronavirusCOVID-19vaccination_-_advice_from_the_JCVI2_December_2020.pdf.

10. CDC. Centers for Disease Control and Prevention. Information about COVID-19 Vaccines for People who Are Pregnant or Breastfeeding Updated March 11 2021 [Available from: https://www.cdc.gov/coronavirus/2019-ncov/vaccines/recommendations/pregnancy.html.

11. JCVI. Joint Committee on Vaccination and Immunisation: advice on priority groups for COVID-19 vaccination, 30 December 2020 2020 [Available from: https://www.gov.uk/government/publications/priority-groups-for-coronavirus-covid-19-vaccination-advice-from-the-jcvi-30-december-2020/joint-committee-on-vaccination-and-immunisation-advice-on-priority-groups-for-covid-19-vaccination-30-december-2020.

12. Shimabukuro TT, Kim SY, Myers TR, Moro PL, Oduyebo T, Panagiotakopoulos L, et al. Preliminary Findings of mRNA Covid-19 Vaccine Safety in Pregnant Persons. New England Journal of Medicine. 2021.

13. PHE. Public Health England. Disparities in the risk and outcomes of COVID-19 2020 [Available from: https://assets.publishing.service.gov.uk/government/uploads/system/uploads/attachment_data/file/891116/disparities_review.pdf.

14. Bell S, Clarke R, Mounier-Jack S, Walker JL, Paterson P. Parents’ and guardians’ views on the acceptability of a future COVID-19 vaccine: A multi-methods study in England. Vaccine. 2020;38(49):7789–98.

15. de Figueiredo A. Sub-national forecasts of COVID-19 vaccine acceptance across the UK: a large-scale cross-sectional spatial modelling study. medRxiv. 2020.

16. Dickerson J, Lockyer B, Moss RH, Endacott C, Kelly B, Bridges S, et al. COVID-19 vaccine hesitancy in an ethnically diverse community: descriptive findings from the Born in Bradford study. 2021.

17. MacKenna B, Curtis H, Morton C, Inglesby P, Walker A, Goldacre B, et al. Trends, regional variation, and clinical characteristics of COVID-19 vaccine recipients: a retrospective cohort study in 23.4 million patients using OpenSAFELY. MedRxiv [Preprint]. 2021.

18. Donaldson B, Jain P, Holder BS, Lindsey B, Regan L, Kampmann B. What determines uptake of pertussis vaccine in pregnancy? A cross sectional survey in an ethnically diverse population of pregnant women in London. Vaccine. 2015;33(43):5822–8.

19. Skirrow H, Holder B, Meinel A, Narh E, Donaldson B, Bosanquet A, et al. Evaluation of a midwife-led, hospital based vaccination service for pregnant women. Human Vaccines & Immunotherapeutics. 2020:1–10.

20. Danchin MH, Costa-Pinto J, Attwell K, Willaby H, Wiley K, Hoq M, et al. Vaccine decision-making begins in pregnancy: Correlation between vaccine concerns, intentions and maternal vaccination with subsequent childhood vaccine uptake. Vaccine. 2018;36(44):6473–9.

21. BBC. Covid: Claims vaccinations harm fertility unfounded 2021 [Available from: https://www.bbc.co.uk/news/health-56012529.

22. Loomba S, de Figueiredo A, Piatek SJ, de Graaf K, Larson HJ. Measuring the impact of COVID-19 vaccine misinformation on vaccination intent in the UK and USA. Nature Human Behaviour. 2021.

23. MaternityVoices. National Maternity Voices Partnership 2021 [Available from: http://nationalmaternityvoices.org.uk/.

24. Clarke V, Braun V. Thematic analysis. Encyclopedia of critical psychology: Springer; 2014. p. 1947–52.

25. Skjefte M, Ngirbabul M, Akeju O, Escudero D, Hernandez-Diaz S, Wyszynski DF, et al. COVID-19 vaccine acceptance among pregnant women and mothers of young children: results of a survey in 16 countries. European journal of epidemiology. 2021;36(2):197–211.

26. Ceulemans M, Foulon V, Panchaud A, Winterfeld U, Pomar L, Lambelet V, et al. Vaccine Willingness and Impact of the COVID-19 Pandemic on Women’s Perinatal Experiences and Practices-A Multinational, Cross-Sectional Study Covering the First Wave of the Pandemic. Int J Environ Res Public Health. 2021;18(7):3367.

27. Goncu Ayhan S, Oluklu D, Atalay A, Menekse Beser D, Tanacan A, Moraloglu Tekin O, et al. COVID-19 vaccine acceptance in pregnant women. Accepted Author Manuscript. International Journal of Gynecology & Obstetrics 2021 https://doiorg/101002/ijgo13713.

28. Campbell H, Edwards A, Letley L, Bedford H, Ramsay M, Yarwood J. Changing attitudes to childhood immunisation in English parents. Vaccine. 2017;35(22):2979–85.

29. SAGE. Factors influencing COVID-19 vaccine uptake among minority ethnic groups. Paper prepared by the ethnicity sub-group of the Scientific Advisory Group for Emergencies (SAGE). Published January. 2021 [Available from: https://www.gov.uk/government/publications/factors-influencing-covid-19-vaccine-uptake-among-minority-ethnic-groups-17-december-2020.

30. Razai MS, Osama T, McKechnie DGJ, Majeed A. Covid-19 vaccine hesitancy among ethnic minority groups. BMJ. 2021;372:513.

31. Heath PT, Culley FJ, Jones CE, Kampmann B, Le Doare K, Nunes MC, et al. Group B streptococcus and respiratory syncytial virus immunisation during pregnancy: a landscape analysis. The Lancet Infectious Diseases. 2017;17(7):e223–e34.

32. Wilson RJ, Paterson P, Jarrett C, Larson HJ. Understanding factors influencing vaccination acceptance during pregnancy globally: a literature review. Vaccine. 2015;33(47):6420–9.

33. Byrne L, Ward C, White JM, Amirthalingam G, Edelstein M. Predictors of coverage of the national maternal pertussis and infant rotavirus vaccination programmes in England. Epidemiology and Infection. 2018;146(2):197–206.

34. McAuslane H, Utsi L, Wensley A, Coole L. Inequalities in maternal pertussis vaccination uptake: a cross-sectional survey of maternity units. Journal of Public Health. 2018;40(1):121–8.

35. Forster AS, Rockliffe L, Chorley AJ, Marlow LAV, Bedford H, Smith SG, et al. Ethnicity-specific factors influencing childhood immunisation decisions among Black and Asian Minority Ethnic groups in the UK: a systematic review of qualitative research. Journal of Epidemiology and Community Health. 2017;71(6):544.

36. RSPH. Royal Society for Public Health. Moving The Needle. Promoting vaccination uptake across the life course.; 2019.

37. Robertson E, Reeve KS, Niedzwiedz CL, Moore J, Blake M, Green M, et al. Predictors of COVID-19 vaccine hesitancy in the UK household longitudinal study. Brain, Behavior, and Immunity. 2021;94:41–50.

38. Bianchi DW, Kaeser L, Cernich AN. Involving Pregnant Individuals in Clinical Research on COVID-19 Vaccines. JAMA. 2021;325(11):1041–2.

39. Kampmann B. Women and children last? Shaking up exclusion criteria for vaccine trials. Nature Medicine. 2021;27(1):8-.

40. Male V. Are COVID-19 vaccines safe in pregnancy? Nature Reviews Immunology. 2021;21(4):200–1.

41. Bardají A, Sevene E, Cutland C, Menéndez C, Omer SB, Aguado T, et al. The need for a global COVID-19 maternal immunisation research plan. The Lancet.

42. IGHI. Institute Global Health Innovation. Covid data hub. Covid-19: Global attitudes towards a COVID-19 vaccine. February 2021 [Available from: https://www.imperial.ac.uk/media/imperial-college/institute-of-global-health-innovation/EMBARGOED-0502.-Feb-21-GlobalVaccineInsights_ICL-YouGov-Covid-19-Behaviour-Tracker_20210301.pdf.

