## Supplementary table 1 for "Women’s views on accepting COVID-19 vaccination during and after pregnancy, and for their babies: A multi-methods study in the UK"

**Supplemental Table 1: Predictors of COVID-19 vaccine acceptance**

| Variable |  | | | | | | | | |
| --- | --- | --- | --- | --- | --- | --- | --- | --- | --- |
|  | **Pregnant** | | | **Non-pregnant** | | | **Baby** | | |
|  | *OR* | *95% CI* | *P value* | *OR* | *95% CI* | *P value* | *OR* | *95% CI* | *P value* |
| Ethnicity |  |  |  |  |  |  |  |  |  |
| *White#* | - | *-* | **-** | - | *-* | **-** | - | *-* | **-** |
| *Black/Black mixed* | 3.98 | *1.97, 8.03* | **<0.005** | 3.91 | *1.92, 7.98* | **<0.005** | 4.21 | *2.09, 8.49* | **<0.005** |
| *Asian/Asian mixed* | 1.36 | *0.81, 2.30* | 0.248 | 1.01 | *0.58, 1.77* | 0.965 | 1.44 | *0.86, 2.42* | 0.171 |
| *Other* | 7.64 | *1.36, 42.96* | **0.021** | 2.00 | *0.42, 9.60* | 0.386 | 2.73 | *0.65, 11.48* | 0.170 |
| Age |  |  |  |  |  |  |  |  |  |
| *Under 25y* | 1.65 | *1.03, 2.64* | **0.038** | 1.64 | *1.01, 2.64* | **0.044** | 1.32 | *0.81, 2.15* | 0.263 |
| *25 – 29y* | 1.21 | *0.93, 1.58* | 0.157 | 1.20 | *0.91, 1.59* | 0.191 | 1.18 | *0.90, 1.54* | 0.237 |
| *30 – 34 y #* | - | *-* | - | - | *-* | - | - | *-* | - |
| *35 – 39 y* | 1.06 | *0.81, 1.40* | 0.650 | 0.77 | *0.57, 1.04* | 0.089 | 0.98 | *0.75, 1.29* | 0.890 |
| *Over 39y* | 1.04 | *0.64, 1.68* | 0.884 | 0.92 | *0.54, 1.56* | 0.758 | 1.13 | *0.69, 1.83* | 0.633 |
| Country |  |  |  |  |  |  |  |  |  |
| *England#* | - | *-* | - | - | *-* | **-** | - | *-* | **-** |
| *Scotland* | 0.72 | *0.41, 1.26* | 0.253 | 0.30 | *0.14, 0.66* | **0.003** | 0.36 | *0.20, 0.66* | **0.001** |
| *Wales* | 1.32 | *0.71, 2.46* | 0.375 | 0.97 | *0.50, 1.88* | 0.932 | 1.06 | *0.57, 1.99* | 0.846 |
| *Northern Ireland* | 1.77 | *0.83, 3.79* | 0.141 | 2.02 | *0.91, 4.50* | 0.085 | 2.32 | *1.08, 5.00* | **0.032** |
| Income |  |  |  |  |  |  |  |  |  |
| *Under £24,999* | 1.77 | *1.18, 2.67* | **0.006** | 2.70 | *1.75, 4.14* | **<0.005** | 1.69 | *1.12, 2.55* | **0.013** |
| *£25,000-£44,999* | 1.57 | *1.13, 2.20* | **0.008** | 1.78 | *1.24, 2.56* | **0.002** | 1.26 | *0.90, 1.77* | 0.178 |
| *£45,000-£64,999* | 1.12 | *0.83, 1.52* | 0.451 | 1.49 | *1.06, 2.08* | **0.020** | 1.02 | *0.77, 1.42* | 0.791 |
| *£65,000-£84,999* | 1.03 | *0.74, 1.43* | 0.850 | 1.16 | *0.81, 1.67* | 0.425 | 0.89 | *0.64, 1.25* | 0.512 |
| *Over £85,000#* | - | *-* | - | - | *-* | - | - | *-* | - |

**OR:** ordinal odds ratio. An OR above 1 indicates a higher likelihood of women giving responses moving from ‘definitely yes’ towards ‘definitely no’ on the Likert scale. **95% CI**: 95% confidence interval. **#** indicates the comparator group in the analysis. **Ethnicity Groups:** **White:** White-British, White-Irish, White-Other, Mixed White/Other; **Black:/Black mixed** Black-British African, Black-British Caribbean, Mixed White-Black Caribbean, Mixed White-Black African, Black-Other; **Asian/Asian mixed:** Asian Indian, Asian-Pakistani, Asian-Bangladeshi, Asian-Other and Chinese, Mixed White-Asian; Mixed: **Other:** Other ethnicity.
