## Supplementary table 2 for "Women’s views on accepting COVID-19 vaccination during and after pregnancy, and for their babies: A multi-methods study in the UK"

### **Supplemental Figure 2: Multivariate analysis of predictors of COVID-19 vaccine acceptance of English residents**

| Variable |  | | | | | | | | |
| --- | --- | --- | --- | --- | --- | --- | --- | --- | --- |
|  | **Pregnant** | | | **Non-pregnant** | | | **Child** | | |
|  | *OR* | *95% CI* | *P value* | *OR* | *95% CI* | *P value* | *OR* | *95% CI* | *P value* |
| Ethnicity |  |  |  |  |  |  |  |  |  |
| *White#* | - | - | - | - | - | - | - | *-* | - |
| *Minority ethnicities* | 2.13 | *1.35, 3.35* | **0.001** | 1.84 | *1.15, 2.95* | **0.011** | 2.25 | *1.43, 3.52* | **<0.005** |
| Region |  |  |  |  |  |  |  |  |  |
| *Greater London#* | - | *-* | - | - | *-* | - | - | *-* | - |
| *East Midlands* | 0.87 | *0.50, 1.51* | 0.616 | 1.09 | *0.60, 2.00* | 0.759 | 1.07 | *0.62, 1.86* | 0.803 |
| *West Midlands* | 1.48 | *0.88, 2.48* | 0.136 | 1.67 | *0.97, 2.88* | 0.066 | 1.49 | *0.89, 2.51* | 0.131 |
| *North East* | 0.62 | *0.33, 1.19* | 0.152 | 0.73 | *0.35, 1.53* | 0.404 | 0.89 | *0.46, 1.73* | 0.738 |
| *North West* | 1.33 | *0.86, 2.05* | 0.199 | 1.38 | *0.86, 2.21* | 0.183 | 1.45 | *0.94, 2.24* | 0.095 |
| *South East* | 1.24 | *0.83, 1.87* | 0.298 | 1.15 | *0.74, 1.80* | 0.529 | 1.26 | *0.84, 1.90* | 0.267 |
| *South West* | 1.04 | *0.62, 1.74* | 0.880 | 1.71 | *1.00, 2.93* | **0.050** | 1.30 | *0.78, 2.17* | 0.308 |
| *Yorkshire, Humber* | 1.03 | *0.67, 1.58* | 0.899 | 1.55 | *0.98, 2.45* | 0.063 | 1.21 | *0.79, 1.88* | 0.373 |
| *East of England* | 1.36 | *0.84, 2.20* | 0.214 | 1.01 | *0.59, 1.72* | 0.980 | 1.36 | *0.84, 2.21* | 0.217 |
| Age |  |  |  |  |  |  |  |  |  |
| *Under 25 y* | 1.08 | *0.64, 1.85* | 0.768 | 0.99 | *0.58, 1.71* | 0.983 | 0.72 | *0.42, 1.25* | 0.248 |
| *25 – 29 y* | 1.11 | *0.83, 1.49* | 0.462 | 1.10 | *0.81, 1.49* | 0.538 | 1.04 | *0.78, 1.39* | 0.776 |
| *30 – 34 y#* | - | *-* | - | - | *-* | - | - | *-* | - |
| *35 – 39 y* | 1.02 | *0.76, 1.37* | 0.916 | 0.77 | *0.56, 1.07* | 0.122 | 0.92 | *0.68, 1.23* | 0.680 |
| *Over 39 y* | 0.93 | *0.56, 1.54* | 0.778 | 0.81 | *0.46, 1.44* | 0.476 | 1.03 | *0.61, 1.73* | 0.613 |
| Income |  |  |  |  |  |  |  |  |  |
| *Under £24,999* | 2.08 | *1.37, 3.15* | **0.001** | 2.47 | *1.59, 3.84* | **<0.005** | 2.26 | *1.49, 3.44* | **<0.005** |
| *£25,000, £44,999* | 1.42 | *0.97, 2.07* | 0.070 | 1.45 | *0.97, 2.18* | 0.071 | 1.23 | *0.84, 1.79* | 0.291 |
| *£45,000, £64,999* | 1.09 | *0.78, 1.53* | 0.613 | 1.35 | *0.93, 1.96* | 0.117 | 0.99 | *0.70, 1.40* | 0.974 |
| *£65,000, £84,999* | 1.00 | *0.71, 1.44* | 0.963 | 1.02 | *0.69, 1.52* | 0.914 | 0.88 | *0.62, 1.26* | 0.481 |
| *Over £85,000#* | - | *-* | - | - | *-* | - | - | *-* | - |

**OR:** ordinal odds ratio. An OR above 1 indicates a higher likelihood of women giving responses moving from ‘definitely yes’ towards ‘definitely no’ on the Likert scale.

**95% CI:** 95% confidence interval.

**#** indicates the comparator group in the analysis.

**Ethnicity Groups:**

**White:** White-British, White-Irish, White-Other.

**Minority ethnicity:** Black-British African, Black-British Caribbean and Black-Other, Asian Indian, Asian-Pakistani, Asian-Bangladeshi, Asian-Other and Chinese, Mixed White-Black Caribbean, Mixed White-Black African, Mixed White-Asian, Mixed White/Other and Other ethnicity.
